# Time Between Viral Loads for People with HIV during the COVID-19 Pandemic

**DOI:** 10.1101/2021.12.19.21268052

**Authors:** Walid G. El-Nahal, Nicola M. Shen, Jeanne C. Keruly, Joyce L. Jones, Anthony T. Fojo, Yukari C. Manabe, Richard D. Moore, Kelly A. Gebo, Geetanjali Chander, Catherine R. Lesko

**Author notes:** **Corresponding Author:** Walid G. El-Nahal. **Alternate Author:** Catherine R. Lesko, **Corresponding and Alternate Author Address:** Johns Hopkins Medicine - Division of Infectious Diseases, 1830 E Monument St, Room 450B, Baltimore, MD 21205, (corresponding), (alternate). Drs. Chander and Lesko contributed equally to this work.

## Abstract

**Background:** During the COVID-19 pandemic, patients experienced significant care disruptions, including lab monitoring. We investigated changes in the time between viral load (VL) checks for people with HIV associated with the pandemic.

**Methods:** This was an observational analysis of VLs of people with HIV in routine care at a large subspecialty clinic. At pandemic onset, the clinic temporarily closed its onsite laboratory. The exposure was time period (time-varying): pre-pandemic (January 1^st^ 2019-March 15^th^, 2020); pandemic lab-closed (March 16^th^-July 12^th^, 2020); and pandemic lab-open (July 13^th^-December 31^st^, 2020). We estimated time from an index VL to a subsequent VL, stratified by whether the index VL was suppressed (≤200 copies/mL). We also calculated cumulative incidence of a non-suppressed VL following a suppressed index VL, and of re-suppression following a loss of viral suppression.

**Results:** Compared to pre-pandemic, hazard ratios for next VL check were: 0.34 (95% CI: 0.30, 0.37, lab-closed) and 0.73 (CI: 0.68, 0.78, lab-open) for suppressed patients; 0.56 (CI: 0.42, 0.79, lab-closed) and 0.92 (95% CI: 0.76, 1.10, lab-open) for non-suppressed patients. The 12-month cumulative incidence of loss of suppression was the same in the pandemic lab-open (4%) and pre-pandemic period (4%). The hazard of re-suppression following loss of suppression was lower during the pandemic lab-open versus the pre-pandemic period (hazard ratio: 0.68, 95% CI: 0.50, 0.92).

**Conclusions:** Early pandemic restrictions and lab closure significantly delayed VL monitoring. Once the lab re-opened, non-suppressed patients resumed normal monitoring. Suppressed patients still had a delay, but no significant loss of suppression.

**Summary:** During the early COVID-19 pandemic, people with HIV experienced disruptions in viral load monitoring due to lab closure and pandemic restrictions. Loosening restrictions resolved delays for non-suppressed, but not suppressed patients. Delays did not significantly increase proportion of non-suppressed patients.

## Introduction

Viral suppression is critical to ending the HIV epidemic,^1^ as patients whose viral load is undetectable are unlikely to transmit infection.^2,3^ Viral suppression with antiretroviral therapy (ART) has also drastically reduced morbidity and mortality rates for people with HIV (PWH).^4,5^ Given the importance of viral suppression, viral load monitoring is recommended frequently at initiation of therapy, then every 3-6 months thereafter depending on the duration of durable suppression.

The COVID-19 pandemic has had a significant impact on the delivery of HIV care, including viral load monitoring. The pandemic precipitated a rapid shift of the bulk of care from in-person to telemedicine.^6^ Social distancing recommendations limited use of public transit during the pandemic,^7^ a barrier to lab collection for patients without access to private transportation.^8^ Surveys suggest Ryan White clinics saw a significant decline in lab monitoring frequency during early months of the pandemic,^9^ with some closing their onsite laboratories as part of social distancing measures. Additionally, early pandemic guidance also advised providers to delay lab monitoring for patients who have been suppressed.^10^

Herein, we aimed to describe time between viral load measurements for people living with HIV during the COVID-19 pandemic compared to pre-pandemic. We stratified analyses by whether the index (i.e., first) viral load in a pair of viral load measurements was suppressed or not, recognizing that the frequency of monitoring depends on prior suppression status even under normal conditions. We also identified groups at risk for longer gaps in viral monitoring during the pandemic. Finally, in secondary analyses, we estimated the cumulative incidence of having a non-suppressed viral load after a suppressed index viral load, and of time to viral load suppression among patients who were unsuppressed at enrollment in the clinic, and among established patients who lost viral suppression during follow-up, comparing pandemic to pre-pandemic periods.

## Methods

### Study Sample

The John G. Bartlett Specialty Practice is a large subspecialty clinic in East Baltimore affiliated with the Johns Hopkins Hospital. The clinic provides comprehensive continuity care to people with HIV or Hepatitis C. The Johns Hopkins HIV Clinical Cohort enrolls clinic patients who consent to share their medical record data. The cohort has been described previously;^11^ data include self-reported age, gender, race, ethnicity, HIV acquisition risk factors, provider encounters, hospital admissions, lab data, prescribed treatment, and clinical diagnoses. The unit of analysis was viral load samples, not individuals, and we included all samples collected between January 1^st^, 2019 and December 31^st^, 2020. In measuring the time between viral loads, each observation consisted of an “index” viral load and the subsequent “outcome” viral load (if a subsequent test was collected before censoring). All index viral loads also served as outcome viral loads for the prior index viral load, with the exception of each individual’s first viral load in the study period. Consequently, individuals who had multiple viral loads collected during the study period could contribute multiple observations to the analysis (Supplemental Figure 1).

### Exposure of Interest

In response to the COVID-19 pandemic, the John G. Bartlett Specialty Practice closed its onsite laboratory and converted most encounters from in-person to telemedicine on March 16^th^, 2020.^12^ Labs for the first 4 months of the pandemic had to be collected offsite. Then on July 12^th^, 2020 the onsite lab reopened. Our exposure was time-varying calendar period (i.e., probability of having a follow-up viral load test was a function of calendar period of follow-up, rather than as a function of calendar period in which the index viral load was collected). The pre-pandemic period was January 1^st^, 2019 to March 15^th^, 2020. The pandemic lab-closed period was March 16^th^, 2020-July 12^th^, 2020 and the pandemic lab-open period was July 13^th^, 2020-December 31^st^, 2020.

### Outcome of Interest

The outcome of interest was the time from each “index” viral load to a subsequent “outcome” viral load.

### Covariates

We were interested in risk factors for longer times between viral load tests and considered the following covariates: age category (20-39 years, 40-59 years, ≥60 years), gender, race, and ethnicity. Recent (within past 6 months) use of alcohol, cocaine or heroin was based on the findings of trained chart abstractors utilizing provider notes, toxicology testing, and treatment referrals, We also included insurance status (private or non-private, where non-private included Ryan White, Medicaid, or uninsured).

### Statistical Analysis

We reported hazard ratios (HR) and 95% confidence intervals (CI) from a Cox proportional hazards model and cumulative incidence curves for time from index viral load to subsequent viral load, comparing each of the two pandemic periods (lab closed and lab open) to the pre-pandemic period as our exposure. Cumulative incidence curves are the complement of the Kaplan-Meier function; we calculated and reported risk at 3, 6, and 12 months from the index viral load. Exposure was time-varying; if no subsequent viral load was collected by the end of a study period, that observation was censored and late-entered into the following period. We administratively censored time at 12 months or the cohort close date, December 31, 2020, whichever came first.

To describe risk factors for delays in viral load monitoring due to the pandemic, we report HRs for hazard of a subsequent viral load associated with time period from a Cox proportional hazards model within strata of each covariate above. We report p-values for an interaction term between each covariate and time period to identify whether the association between viral load monitoring and time period was different by patient covariate.

Recognizing that the frequency of viral load monitoring differs based on a patient’s suppression status, we stratified all analyses by whether the index viral load in each observation was suppressed (≤200 copies/ml) or not. The suppressed analysis was adjusted for duration of time a patient had previously been suppressed for, recognizing that recently suppressed patients may be monitored more frequently.

### Secondary Analyses: Discordant Viral Loads

Our primary analysis was stratified by the index viral load, irrespective of whether the subsequent viral load was suppressed or not. In secondary analyses we aimed to investigate discordant viral load pairs. The goal of these analyses was to provide insight into (1) loss of suppression during the pandemic and (2) the time to suppression during the pandemic.

### Suppressed to Non-suppressed

For this analysis we restricted to observations with a suppressed index viral load. The outcome of interest was time to the subsequent viral load and whether it was non-suppressed. We treated subsequent viral loads that were suppressed as competing events. We report the cumulative incidence of non-suppression for each calendar period (estimated using the Aalen-Johansen estimator) and report sub-distribution hazard ratios for time to non-suppression by period estimated from Fine and Gray models.

### Non-suppressed to Suppressed

For this analysis, we restricted to observations with a non-suppressed index viral load. We stratified these into two clinically distinct groups: viral loads from (i) patients who were non-suppressed at enrollment and (ii) established patients who became non-suppressed after having been previously suppressed. Rather than time to subsequent viral load, we were interested in time to (i) initial suppression or (ii) re-suppression. The outcome of interest was time to first subsequent suppressed viral load (≤200 copies/ml) (interim viral loads that were not yet suppressed were disregarded). We report the cumulative incidence of suppression for each calendar period and report sub-distribution hazard ratios for time to suppression by period estimated from Cox proportional hazards models.

## Results

### Viral Load Observations

There were 5,498 (70.9%) suppressed index viral loads pre-pandemic (1/1/2019-3/15/2020), 618 (7.9%) during the pandemic lab-closed period (3/16/2020-7/12/2020), and 1,644 (21.2%) during the pandemic lab-open period (7/13/2020-12/31/2020); 3,950 people contributed ≥1 observation to this analysis. There were 750 (73.2%) non-suppressed index viral loads pre-pandemic, 84 (8.2%) during the pandemic lab-closed period, and 191 (18.6%) during the pandemic lab-open period; 601 people contributed ≥1 observation to this analysis. Viral loads were contributed by people that were predominantly age 40-59, male, Black and non-Hispanic (Table 1).

**Table 1:** Patient Characteristics for Viral Load Samples by Time Period for Suppressed and Non-Suppressed Index Viral Loads.

|  | Non-Suppressed |  |  | Suppressed<sup>a</sup> |  |  |
| --- | --- | --- | --- | --- | --- | --- |
|  | Pre-Pandemic | Pandemic, Lab Closed | Pandemic, Lab Open | Pre-Pandemic | Pandemic, Lab Closed | Pandemic, Lab Open |
| **Individuals** | n = 403 | n = 63 | n = 135 | n = 2,065 | n = 565 | n = 1,320 |
| **Person-Time (Days)** | 59,965 | 19,050 | 24,325 | 650,321 | 220,311 | 296,279 |
| **Viral Loads Collected** | 750 | 84 | 191 | 5,498 | 618 | 1,644 |
| **Age** |  |  |  |  |  |  |
| Age 20-39 | 247 (33%) | 30 (36%) | 63 (33%) | 820 (15%) | 69 (11%) | 206 (13%) |
| Age 40-59 | 352 (47%) | 34 (40%) | 85 (45%) | 2,753 (50%) | 323 (52%) | 798 (49%) |
| Age 60+ | 151 (20%) | 20 (24%) | 41 (23%) | 1,925 (35%) | 226 (37%) | 640 (39%) |
| **Male** | 454 (61%) | 51 (61%) | 110 (58%) | 3,516 (64%) | 395 (64%) | 1,029 (63%) |
| **Race** |  |  |  |  |  |  |
| Black | 649 (87%) | 72 (86%) | 164 (86%) | 4,313 (78%) | 453 (73%) | 1,266 (77%) |
| White | 79 (11%) | 11 (13%) | 21 (11%) | 967 (18%) | 142 (23%) | 218 (19%) |
| Other | 22 (3%) | 1 (1%) | 3 (3%) | 218 (4%) | 23 (4%) | 60 (4%) |
| **Hispanic** | 12 (2%) | 1 (1%) | 5 (3%) | 146 (3%) | 16 (3%) | 46 (3%) |
| **Recent Smoking<sup>b</sup>** |  |  |  |  |  |  |
| No | 219 (45%) | 27 (50%) | 61 (51%) | 2,945 (65%) | 331 (69%) | 848 (68%) |
| Yes | 266 (55%) | 27 (50%) | 58 (49%) | 1,616 (35%) | 151 (31%) | 392 (32%) |
| Missing | 265 | 30 | 72 | 937 | 136 | 404 |
| **Recent Alcohol<sup>b</sup>** |  |  |  |  |  |  |
| No | 380 (78%) | 43 (80%) | 100 (83%) | 4,069 (90%) | 435 (90%) | 1,135 (92%) |
| Yes | 105 (22%) | 11 (20%) | 21 (17%) | 480 (10%) | 48 (10%) | 105 (8%) |
| Missing | 265 | 30 | 70 | 953 | 135 | 404 |
| **Recent Cocaine/Heroin<sup>b</sup>** |  |  |  |  |  |  |
| No | 372 (77%) | 44 (81%) | 103 (85%) | 4,065 (89%) | 449 (93%) | 1,144 (92%) |
| Yes | 112 (23%) | 10 (19%) | 18 (15%) | 480 (11%) | 35 (7%) | 98 (8%) |
| Missing | 265 | 30 | 70 | 953 | 134 | 402 |
| **Insurance<sup>c</sup>** |  |  |  |  |  |  |
| Public | 392 (52%) | 40 (48%) | 99 (52%) | 1,807 (33%) | 168 (28%) | 511 (31%) |
| Private | 355 (48%) | 43 (52%) | 92 (48%) | 3,647 (67%) | 442 (72%) | 1,119 (69%) |
| Missing | 3 | 1 | 0 | 44 | 8 | 14 |
| **Months Suppressed<sup>d</sup>** | 0 | 0 | 0 | 34 (8,60) | 40 (8, 61) | 38 (9, 65) |

<sup>a</sup> Suppressed if Viral Load ≤200 copies/mL on most recent check.
<sup>b</sup> Smoking, Alcohol, Cocaine and Heroin use were obtained from medical record review of provider notes, toxicology screens and treatment referrals, conducted every 6 months by trained abstractors. Data were restricted to abstractions in the year prior to the viral load.
<sup>c</sup> Non-Private Insurance defined as covered by Medicaid, Ryan White, or Uninsured.
<sup>d</sup> Median (IQR)

### Time to Next Viral Load for Suppressed Patients

Pre-pandemic, the proportion of suppressed viral loads that had a follow-up viral load by 12 months, was 91%. Under pandemic lab-closed conditions, the proportion was 59%, and under pandemic lab-open conditions, it was 87% (Figure 1). Compared to pre-pandemic, the HRs for subsequent viral load collection were 0.34 (95% CI: 0.30, 0.37) during the pandemic lab-closed period and 0.73 (95% CI: 0.68, 0.78) during the pandemic lab-open period (Table 2). The hazard for viral load monitoring during the pandemic lab-open period relative to pre-pandemic was significantly lower for patients who were Black (HR: 0.71, 95% CI: 0.66, 0.76) compared to White (HR: 0.85, 95% CI: 0.74, 0.98), interaction p-value=0.02; patients who had recent cocaine or heroin use (HR: 0.56, 95% CI: 0.43, 0.73) compared to no recent substance use (HR: 0.76, 95% CI: 0.71, 0.82), interaction p-value=0.03; and patients who had non-private insurance (HR: 0.65, 95% CI: 0.58, 0.74) compared to patients with private insurance (HR: 0.77, 95% CI: 0.71, 0.83), interaction p-value=0.03.

**Table 2:** Hazard Ratios for Time Between Viral Loads Among Those with a <u>Non-Suppressed</u> and <u>Suppressed</u> Index Viral Load, Comparing Pandemic Periods to Pre-pandemic.

|  | Pre-Pandemic | Pandemic, Lab Closed |  |  |  | Pandemic, Lab Open |  |  |  |
| --- | --- | --- | --- | --- | --- | --- | --- | --- | --- |
|  |  | Non-Suppressed Hazard Ratio | Interaction p-value<sup>d</sup> | Suppressed Hazard Ratio | Interaction p-value<sup>d</sup> | Non-Suppressed Hazard Ratio | Interaction p-value<sup>d</sup> | Suppressed Hazard Ratio | Interaction p-value<sup>d</sup> |
| **All Viral Loads<sup>a</sup>** | 1.0 | 0.57 (0.45, 0.72) | n/a | 0.34 (0.31, 0.37) | n/a | 0.92 (0.77, 1.09) | n/a | 0.73 (0.68, 0.78) | n/a |
| **Age** |  |  |  |  |  |  |  |  |  |
| Age 20-39 | 1.0 | 0.47 (0.31, 0.71) | REF | 0.24 (0.19, 0.32) | REF | 0.83 (0.61, 1.12) | REF | 0.62 (0.52, 0.74) | REF |
| Age 40-59 | 1.0 | 0.63 (0.45, 0.89) | 0.29 | 0.37 (0.33, 0.42) | 0.01 | 0.92 (0.72, 1.20) | 0.57 | 0.73 (0.66, 0.79) | 0.11 |
| Age 60+ | 1.0 | 0.61 (0.38, 1.00) | 0.42 | 0.33 (0.29, 0.39) | 0.05 | 1.04 (0.73, 1.48) | 0.34 | 0.78 (0.71, 0.87) | 0.05 |
| **Gender** |  |  |  |  |  |  |  |  |  |
| Female | 1.0 | 0.59 (0.42, 0.84) | REF | 0.34 (0.29, 0.39) | REF | 1.1 (0.78, 1.32) | REF | 0.79 (0.71, 0.88) | REF |
| Male | 1.0 | 0.56 (0.41, 0.75) | 0.79 | 0.34 (0.30, 0.38) | 0.97 | 0.86 (0.68, 1.08) | 0.34 | 0.70 (0.64, 0.76) | 0.06 |
| **Race** |  |  |  |  |  |  |  |  |  |
| Black | 1.0 | 0.52 (0.40, 0.68) | REF | 0.31 (0.28, 0.34) | REF | 0.91 (0.75, 1.10) | REF | 0.71 (0.66, 0.76) | REF |
| White | 1.0 | 0.85 (0.48, 1.50) | 0.12 | 0.48 (0.40, 0.59) | <0.01 | 0.87 (0.51, 1.47) | 0.87 | 0.85 (0.74, 0.99) | 0.02 |
| Other | 1.0 | 0.80 (0.23, 2.78) | 0.52 | 0.32 (0.20, 0.51) | 0.92 | 1.23 (0.44, 3.42) | 0.57 | 0.65 (0.47, 0.90) | 0.62 |
| **Ethnicity** |  |  |  |  |  |  |  |  |  |
| Non-Hispanic | 1.0 | 0.56 (0.44, 0.71) | REF | 0.34 (0.31, 0.37) | REF | 0.90 (0.75, 1.08) | REF | 0.73 (0.68, 0.78) | REF |
| Hispanic | 1.0 | 1.59 (0.40, 6.36) | 0.15 | 0.30 (0.16, 0.54) | 0.67 | 2.37 (0.67, 8.40) | 0.14 | 0.74 (0.51, 1.08) | 0.94 |
| **Substance Use<sup>b</sup>** |  |  |  |  |  |  |  |  |  |
| No Recent Smoking | 1.0 | 0.52 (0.35, 0.78) | REF | 0.35 (0.31, 0.40) | REF | 0.85 (0.62, 1.17) | REF | 0.79 (0.73, 0.87) | REF |
| Recent Smoking | 1.0 | 0.50 (0.32, 0.76) | 0.88 | 0.27 (0.23, 0.33) | 0.02 | 1.00 (0.75, 1.34) | 0.44 | 0.66 (0.58, 0.74) | 0.01 |
| No Recent Alcohol Use | 1.0 | 0.51 (0.37, 0.72) | REF | 0.32 (0.29, 0.36) | REF | 0.96 (0.75, 1.22) | REF | 0.75 (0.69, 0.81) | REF |
| Recent Alcohol Use | 1.0 | 0.52 (0.27, 0.98) | 0.98 | 0.35 (0.26, 0.47) | 0.65 | 0.89 (0.56, 1.39) | 0.76 | 0.66 (0.53, 0.83) | 0.30 |
| No Recent Cocaine/Heroin | 1.0 | 0.51 (0.37, 0.71) | REF | 0.34 (0.30, 0.38) | REF | 0.91 (0.72, 1.15) | REF | 0.76 (0.71, 0.82) | REF |
| Recent Cocaine or Heroin | 1.0 | 0.50 (0.26, 0.97) | 0.94 | 0.23 (0.16, 0.33) | 0.04 | 0.98 (0.60, 1.58) | 0.79 | 0.56 (0.44, 0.71) | 0.03 |
| **Insurance<sup>c</sup>** |  |  |  |  |  |  |  |  |  |
| Private | 1.0 | 0.56 (0.40, 0.77) | REF | 0.26 (0.22, 0.31) | REF | 0.92 (0.72, 1.17) | REF | 0.65 (0.58, 0.73) | REF |
| Non-Private | 1.0 | 0.58 (0.42, 0.81) | 0.85 | 0.37 (0.33, 0.41) | <0.01 | 0.90 (0.70, 1.16) | 0.91 | 0.77 (0.71, 0.83) | 0.03 |

<sup>a</sup> Among Non-Suppressed patients: Index Viral Load > 200 copies/mL. Among Suppressed patients: Index Viral Load ≤200 copies/mL.
Suppressed analysis adjusted for duration of suppression prior to index viral load.
<sup>b</sup> Smoking, Alcohol, Cocaine and Heroin use were obtained from medical record review of provider notes, toxicology screens and treatment referrals, conducted every 6 months by trained abstractors. Data were restricted to abstractions in the year prior to the viral load.
<sup>c</sup> Non-Private Insurance defined as covered by Medicaid, Ryan White, or Uninsured.
<sup>d</sup> P-value for the interaction between patient characteristic and the time period, to determine if there were statistically significant differences in hazard across subgroups.

**Figure 1.**
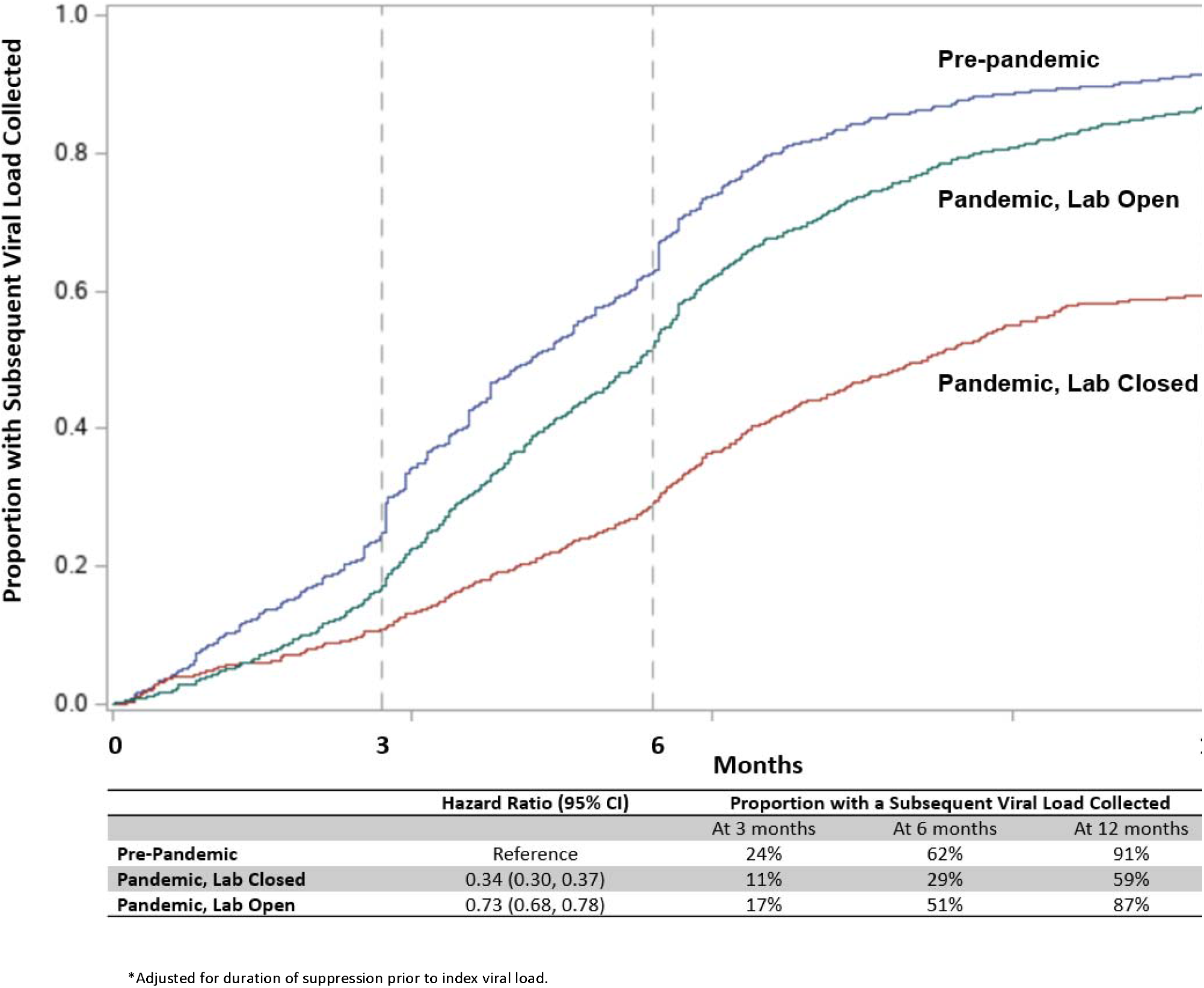
Kaplan-Meier Curve of Time from Suppressed Viral Load to Subsequent Viral Load.

### Time to Next Viral Load for Non-suppressed Patients

Pre-pandemic, the proportion of non-suppressed viral loads that had a follow-up viral load by 12 months, was 90%. Under pandemic lab-closed conditions, the proportion was 75%, while under pandemic lab-open conditions, it was 88% (Figure 2). Compared to pre-pandemic, the HRs for subsequent viral load collection were 0.56 (95% CI: 0.42, 0.79) during the pandemic lab-closed period and 0.92 (95% CI: 0.76, 1.10) during the pandemic lab-open period (Table 2). Among non-suppressed patients, once the lab re-opened, there were no statistically significant differences (compared to pre-pandemic) in time between viral loads across any patient characteristics.

**Figure 2.**
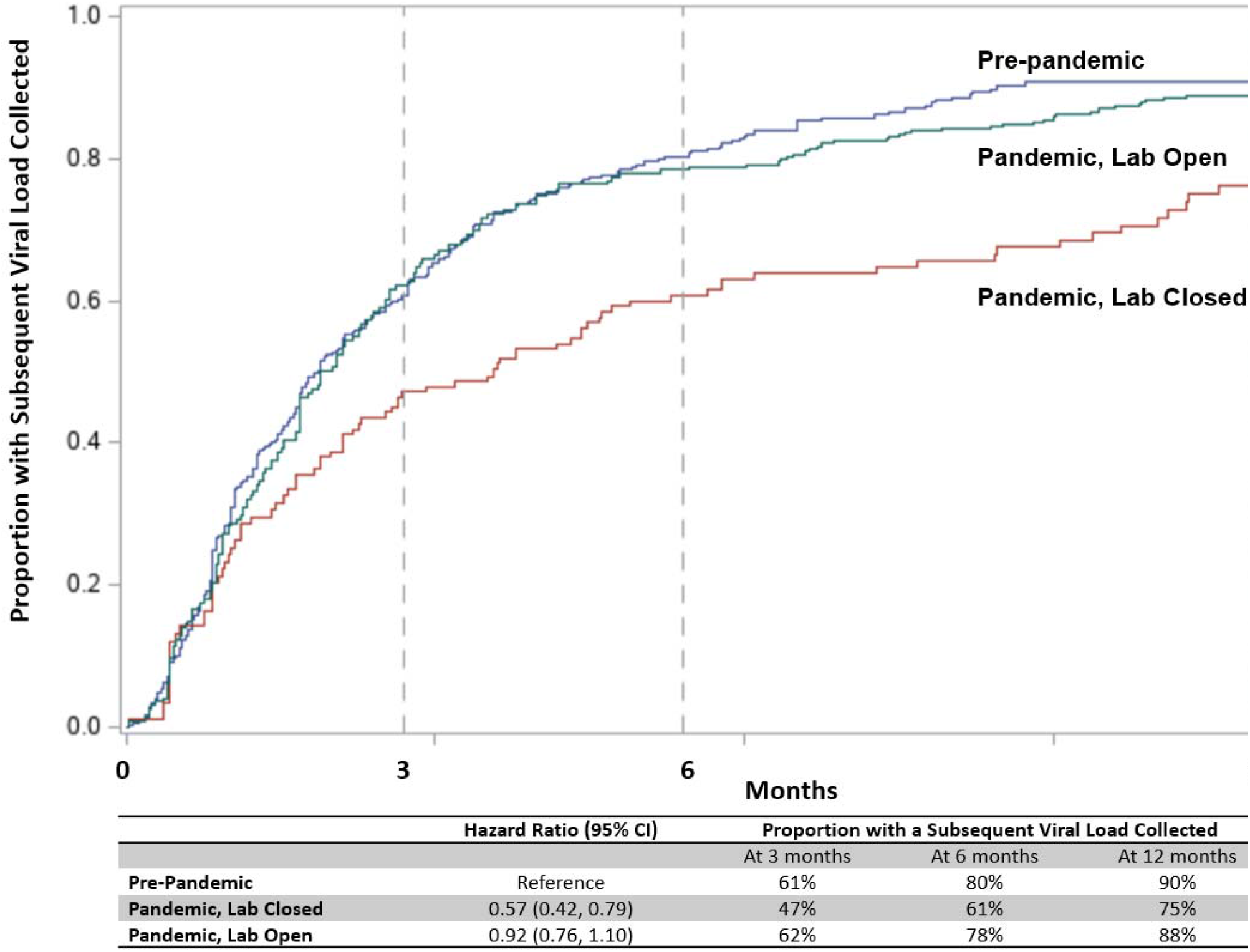
Kaplan-Meier Curve of Time from Non-Suppressed Viral Load to Subsequent Viral Load.

### Loss of Viral Suppression

Following index viral loads that were suppressed, the 12-month cumulative incidence of a subsequent viral loads that was not suppressed was 4% pre-pandemic, 2% under pandemic lab-closed conditions, and 4% under the pandemic lab-open conditions (Supplemental Figure 2). The corresponding hazard ratios compared to pre-pandemic were 0.37 (95% CI: 0.33, 0.41) and 1.04 (95% CI: 0.98, 1.10) respectively.

### Time from Non-Suppressed to (i) Initial Suppression or (ii) Re-Suppression

There were 70 patients who enrolled into the cohort as non-suppressed. The cumulative incidence of initial suppression at 12 months was 66% pre-pandemic, 15% under pandemic lab-closed conditions, and 74% under pandemic lab-open conditions (Supplemental Figure 3). The corresponding HRs for initial suppression compared to pre-pandemic were 0.18 (95% CI: 0.02, 1.29) for pandemic lab-closed conditions and 1.37 (95% CI: 0.61, 3.11) for pandemic lab-open conditions. Among established previously suppressed patients, there were 608 instances of loss of suppression across 352 individuals. The 12-month cumulative incidence of re-suppression was 85% pre-pandemic, 46% under pandemic lab-closed conditions, and 70% under pandemic lab-open conditions (Supplemental Figure 4). The corresponding HRs for re-suppression compared to pre-pandemic were 0.38 (95% CI: 0.25, 0.59) for pandemic lab-closed conditions and 0.68 (95% CI: 0.50, 0.92) for pandemic lab-open conditions.

## Discussion

At the onset of the pandemic, when the onsite lab was closed and pandemic restrictions were implemented, the probability of viral load monitoring was drastically reduced. Once the onsite lab reopened and restrictions loosened, non-suppressed viral loads were re-checked at approximately the same rate as pre-pandemic. For suppressed patients, the rate of re-checking viral loads was lower during the pandemic compared to pre-pandemic. The reduction in viral load monitoring of suppressed patients during the pandemic lab-open period compared to the pre-pandemic period was greatest for patients who were Black, had non-private insurance, or had recent substance use.

Longer time between viral loads during the early pandemic is unsurprising. With closure of the onsite lab, patients were reliant on third party commercial laboratories. These off-site labs were heavily strained by increasing COVID-19 testing demands during the pandemic, while limited by reduced in-person staffing as a means of social distancing, on top of pre-existing lab staffing shortages.^13,14^ Beyond lab-related issues, patients’ ability to reach commercial labs early in the pandemic may have been hindered by disruptions to public transportation during that period.^7^ Additionally, patients often forwent care during this early pandemic period for reasons including anxiety about contracting SARS-CoV-2, financial concerns related to job security during the pandemic, and insurance loss from job turnover during the recession.^15^ The onsite lab re-opening coincided somewhat with a better understanding of appropriate COVID-19 precautions and a gradual re-opening of other services (e.g., public transportation and more in-person clinical visits) and thus we might expect to see the associated shortening time between viral loads that was observed. However, our results may also suggest the importance of access to an onsite laboratory for our patients; certainly, when attending an in-person clinic visit, being able to get labs drawn onsite eliminated the need for additional travel and associated risk of COVID-19 exposure.

Once the lab reopened later in the pandemic, the duration between viral load checks shortened to pre-pandemic levels for patients who were not suppressed, but not for patients who are suppressed. The cause of the difference is uncertain. Informally, providers reported being more aggressive about getting viral load testing for patients who were not suppressed. Conversely, for patients who were known to be suppressed, interim guidance early in the pandemic suggested postponing lab monitoring to the extent possible for patients in otherwise stable health to mitigate risk of COVID-19 exposure.^10^

The longer duration between viral loads for suppressed patients during the pandemic was more prominent among patients who were Black, had non-private insurance, or had a history of substance use disorder. These groups are historically at higher risk for viral non-suppression but also are at higher risk for severe COVID-19, thus while this difference could be interpreted as a disparity, it could also be interpreted as the result of efforts to limit COVID-19 exposure.

Despite the prolonged time between viral loads for suppressed patients during the pandemic, the cumulative incidence of loss of suppression was minimal. While this is reassuring, and may suggest that less frequent lab monitoring of suppressed patients is viable, it is crucial to note that our analysis was based on observed viral load values during a period when viral load monitoring was less frequent. We surely missed instances of non-suppressed viral loads that were not measured; the magnitude of this bias is unknown. Additionally, these findings apply to a unique period in patient care, the COVID-19 pandemic, and whether they apply under “normal” conditions merits further study.

The time to observed re-suppression was significantly impacted by the COVID-19 pandemic. This may be an artifact. Remote delivery of care through telemedicine and delayed lab monitoring might mean that patients resuppressed their viral load at the same rate pre-pandemic and during the pandemic, but we were not able to detect that re-suppression as quickly during the pandemic. However, we hypothesize that a more likely explanation is that during the pandemic, the rate of re-suppression was lower due to forgone care (that may have included ART adherence counseling) due to anxiety about COVID-19 exposure, financial insecurity, or loss of insurance during the pandemic recession; barriers to picking up anti-retroviral therapy (ART); and lower ART adherence due to increased anxiety, depression, substance use, or other factors associated with the COVID-19 pandemic. Our clinic worked to mitigate some of these barriers by offering ART delivery during the pandemic, telemedicine clinical visits, and telemedicine psychiatry visits.

Several limitations are worth noting in this analysis. Viral load monitoring requires the provider order a lab test and the patient get the lab test, and it is unclear from our findings alone whether delays in viral load monitoring were most influenced by less provider ordering or lower patient follow-through. In our analysis of loss of suppression and time to re-suppression, we are limited to viral loads that are observed. We may have underestimated the risk of both outcomes. Additionally, all viral load measurements apply only to a point in time, patients’ viral load between checks is unknown. Thus, we may have underestimated the probability of viral non-suppression due to loss of suppression (and subsequent re-suppression) that occurred during a prolonged, unmonitored period. Our analysis of time to initial suppression was limited by a very small sample size, reflected in wide confidence. Finally, the data were collected during the pandemic, limiting their generalizability and whether they should be used to inform monitoring frequency guidelines moving forward.

## Conclusion

In this study, the closure of onsite labs amid early pandemic restrictions resulted in less frequent monitoring of viral loads for both suppressed and non-suppressed patients. Once the lab re-opened and restrictions loosened, frequency of viral load monitoring for non-suppressed patients returned to pre-pandemic levels. Suppressed patients however continued to be monitored less frequently, with some subgroups affected more than others. The longer monitoring interval does not appear to have resulted in significant loss of suppression. However, patients who did experience a loss of viral suppression took longer to resuppress their viral load during the pandemic.

## Data Availability

All data produced in the present study are available upon reasonable request to the authors.

## Acknowledgements & Funding

This work was supported by grants from the National Institutes of Health [T32 AI007291, K24 AA027483, K01 AA028193, K08 MH118094, U01 DA036935 and P30 AI094189]. The content is solely the responsibility of the authors and does not necessarily represent the official views of the National Institutes of Health.

## Conflicts of Interest

The authors have no conflicts to declare.

## Notes

### Competing Interest Statement

The authors have declared no competing interest.

### Author Declarations

Johns Hopkins Medicine Institutional Review Board gave ethical approval for this work.

