## Supplemental Materials for "Time Between Viral Loads for People with HIV during the COVID-19 Pandemic"

**Supplementary Materials**

**Supplemental Figure 1 – Viral Load Pair Observation Contributions to each Time Period**

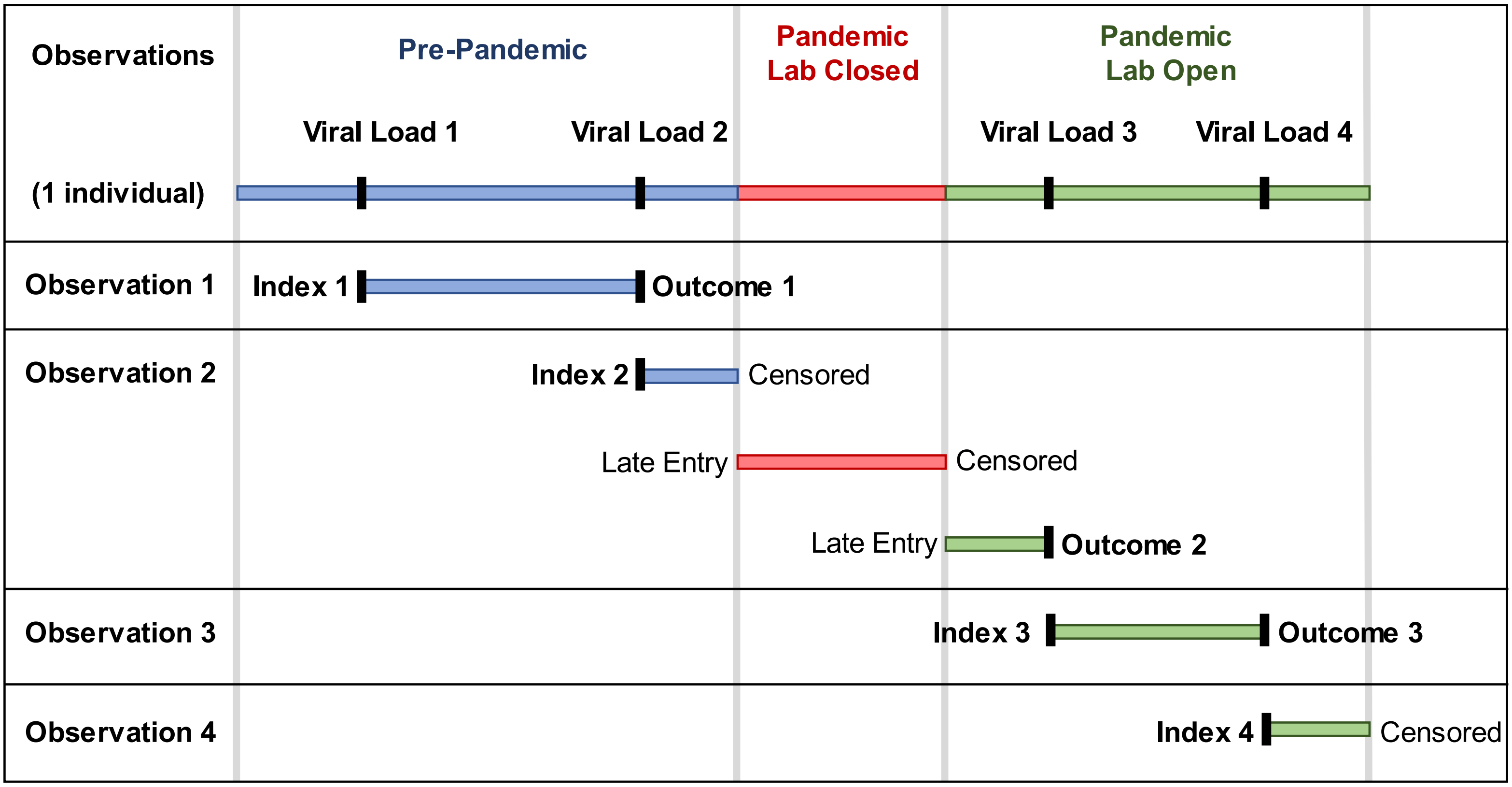

**Supplemental Figure 2 - Kaplan-Meier Curve of Suppressed to Subsequent Non-Suppressed Viral Load**

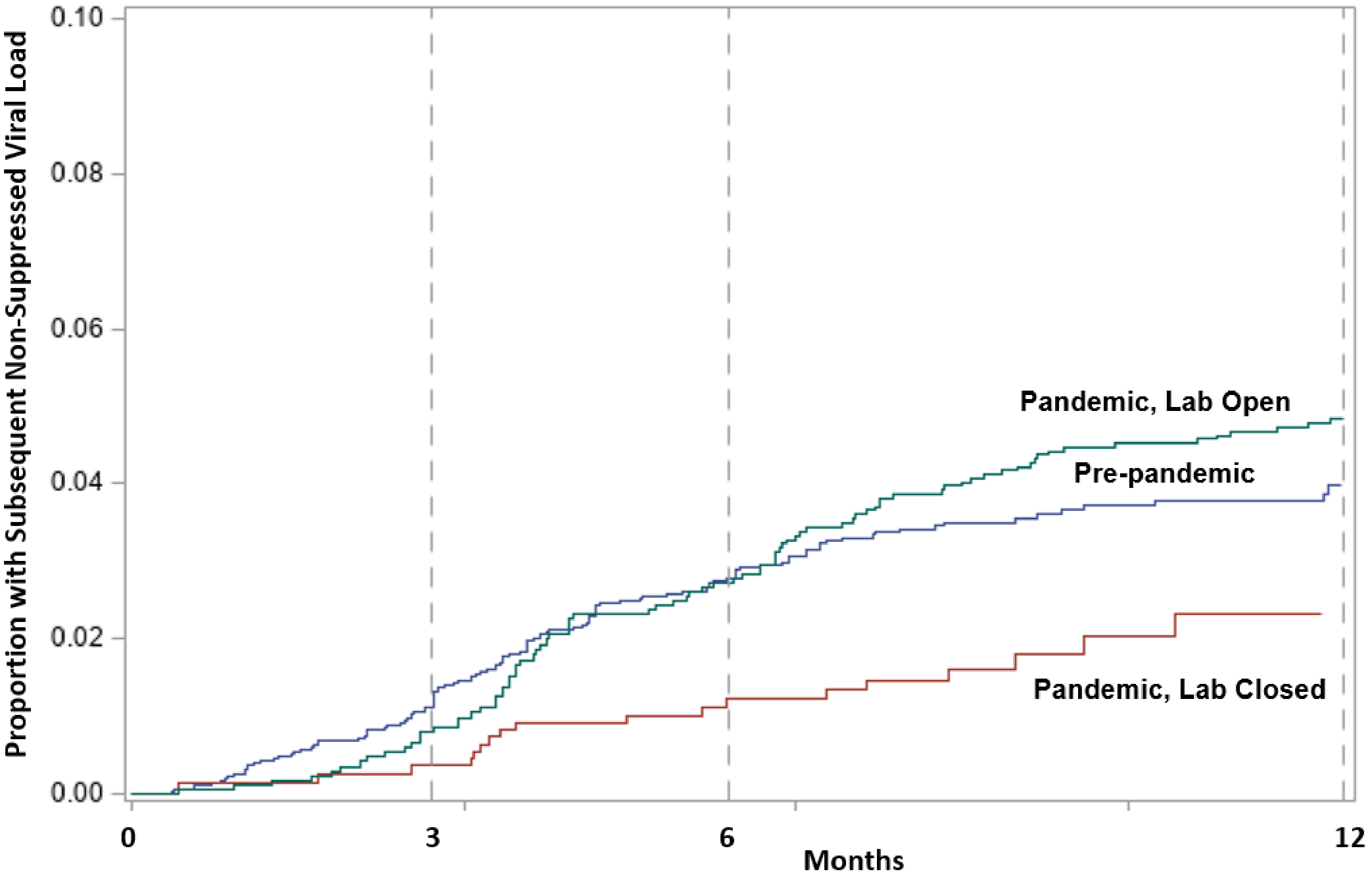


| | Hazard Ratio (95% CI) | Proportion with a Subsequent Non-suppressed Viral Load | | |
|---|---|---|---|---|
| | | At 3 months | At 6 months | At 12 months |
| **Pre-Pandemic** | Reference | 1% | 3% | 4% |
| **Pandemic, Lab Closed** | 0.37 (0.33, 0.41) | 0% | 1% | 2% |
| **Pandemic, Lab Open** | 1.04 (0.98, 1.10) | 1% | 2% | 4% |

**Supplemental Figure 3 - Kaplan-Meier Curve from Non-Suppressed at Enrollment to Initial Suppression**

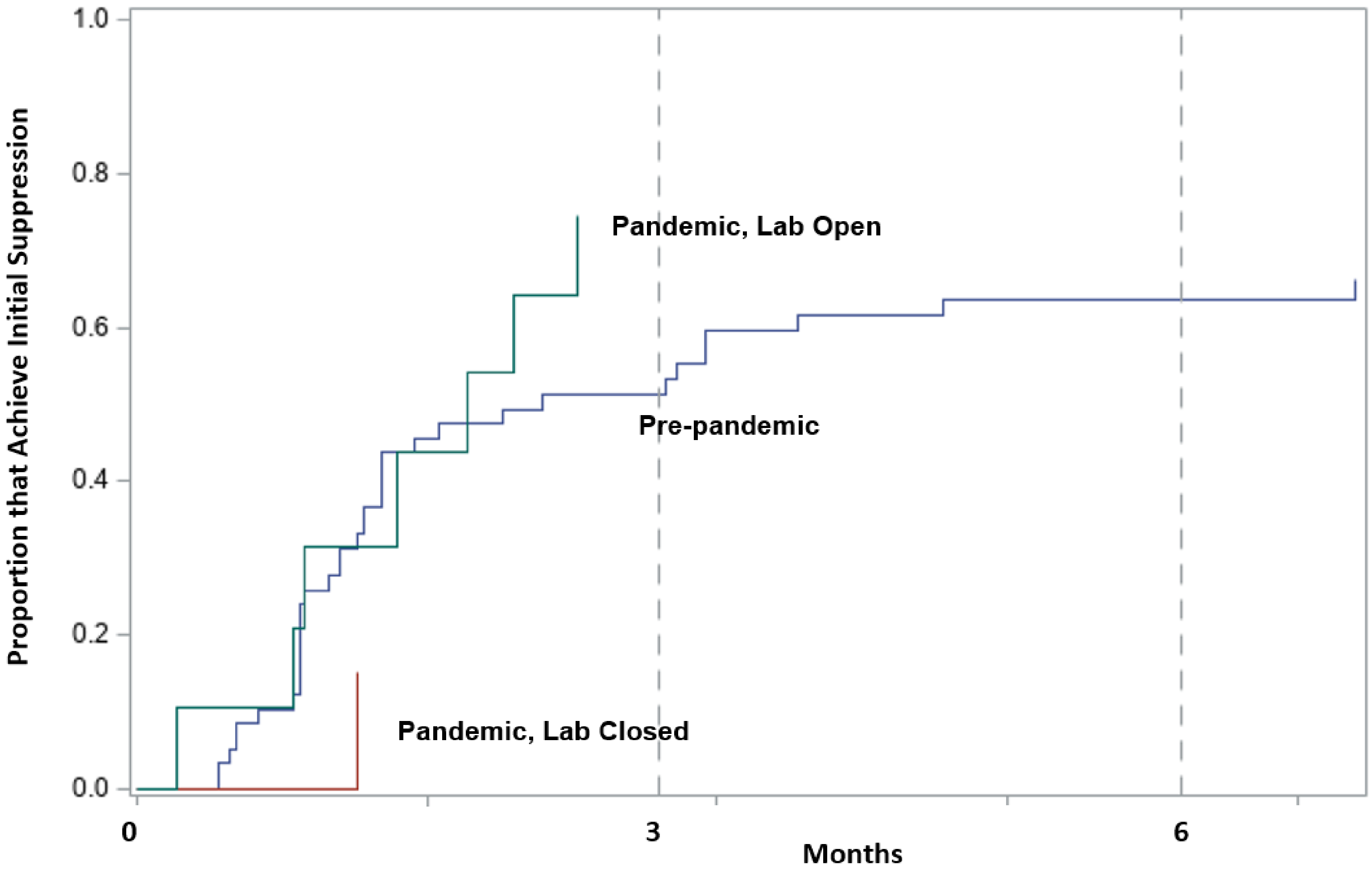


| | Hazard Ratio (95% CI) | Proportion that Achieve Initial Suppression | | |
|---|---|---|---|---|
| | | At 3 months | At 6 months | At 12 months |
| **Pre-Pandemic** | Reference | 51% | 64% | 66% |
| **Pandemic, Lab Closed** | 0.18 (0.02, 1.29) | 15% | 15% | 15% |
| **Pandemic, Lab Open** | 1.37 (0.61, 3.11) | 74% | 74% | 74% |

**Supplemental Figure 4 - Kaplan-Meier Curve from Loss of Suppression to Re-Suppression**

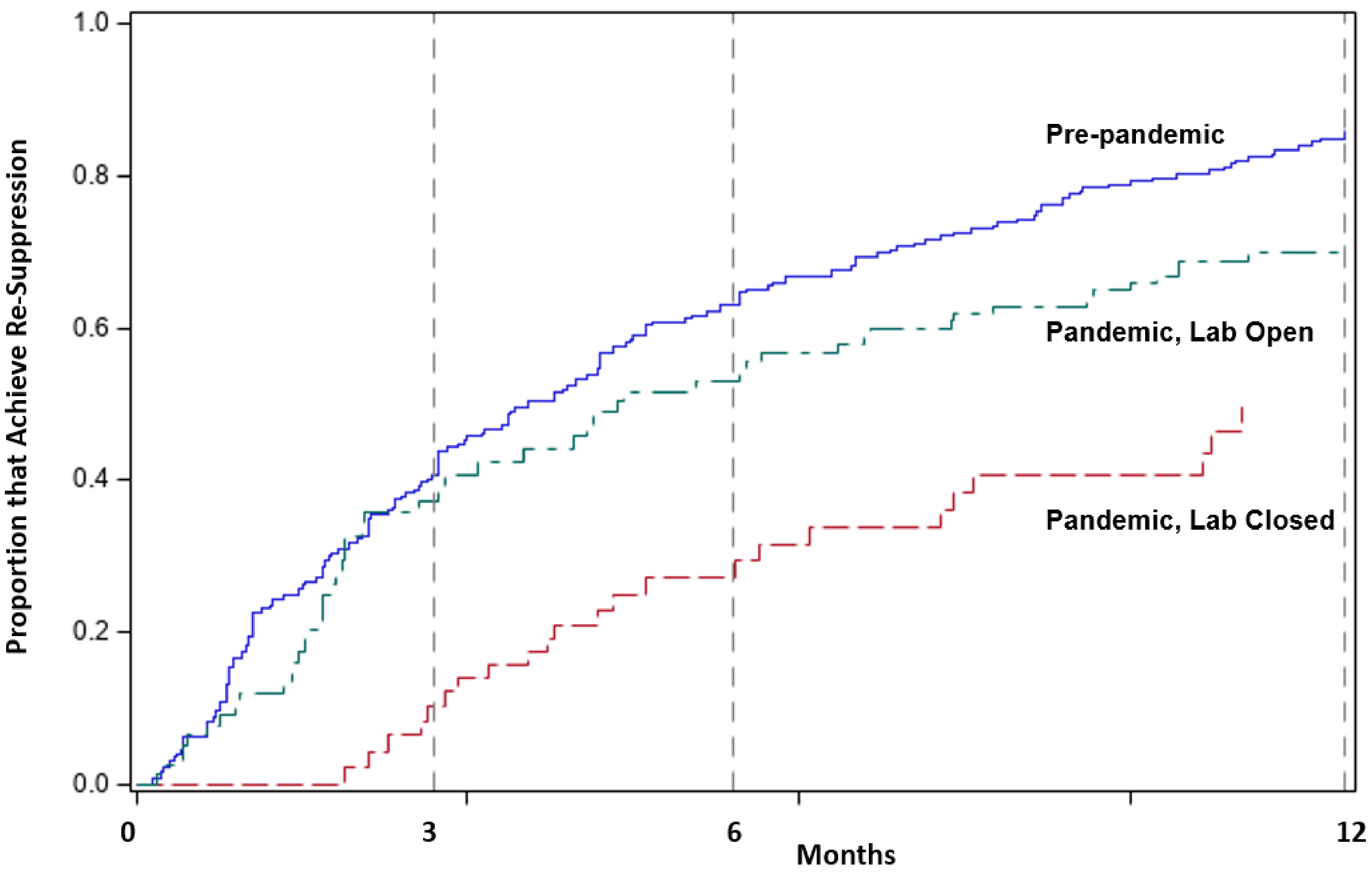


| | Hazard Ratio (95% CI) | Proportion that Achieve Re-Suppression | | |
|---|---|---|---|---|
| | | At 3 months | At 6 months | At 12 months |
| **Pre-Pandemic** | Reference | 41% | 63% | 85% |
| **Pandemic, Lab Closed** | 0.38 (0.25, 0.59) | 10% | 27% | 46% |
| **Pandemic, Lab Open** | 0.68 (0.50, 0.92) | 37% | 53% | 70% |
